# Evaluating Linkage Approaches for Address-Level Socioenvironmental Exposure Assessment

**DOI:** 10.64898/2025.12.02.25341378

**Authors:** Carson S. Hartlage, Erika Rasnick Manning, Cole Brokamp

## Abstract

Accurate linkage of addresses to parcel-level data is essential for hyperlocal environmental exposure assessment, yet the performance of methods, including their impact on exposure misclassification and bias, remains poorly characterized. Using a gold standard match of 853,255 National Address Database records to authoritative datasets from Hamilton and Franklin Counties, Ohio, we evaluated address tag fuzzy matching and geocoding-based (geomatching) approaches on accuracy of linked parcel identifier and parcel market total value and usage type. Address tag fuzzy matching achieved 100% agreement; address point geomatching performed moderately well (65.1% - 76.1%), and street range geomatching performed poorly (7.2% - 59.2%). Poorer agreement was more common in neighborhoods with higher address densities and more community material deprivation, highlighting potential for differential misclassification of exposure assessment. These findings emphasize a need for precise, scalable, and standardized linkage approaches to support valid address- and parcel-level exposure assessment in clinical and population health research.

## Introduction

Addresses are foundational spatial identifiers through which individuals and institutions operationalize precise geographic location. In clinical and public health settings, addresses support surveillance, risk stratification, and equity monitoring by enabling linkage of patient data to environmental and socioeconomic indicators.^1^ Within electronic health records, accurate geocoding enables evaluation of spatial clusters of risk,^2^ neighborhood-level inequities,^3^ and how residential mobility shapes health trajectories over time.^4^

In environmental epidemiology, accurate addresses are essential for assigning exposures that vary over space and time, such as air pollution, noise, temperature, or greenspace. Geocoding, the process of transforming address text into spatial coordinates, underpins this linkage between individuals and environmental data. Street-range geocoding is a commonly used approach that relies on interpolating locations based on the range of street numbers on a street centerline; while this method is adequate for neighborhood or census-level analyses, its positional uncertainty makes it poorly suited for linking to smaller geographic units.^5,6^

Parcel data, such as home value, structure type, land use class, and building condition for legal units of land, are often made publicly available by county auditors and provide a valuable source of information for researchers seeking to investigate the impacts of socioeconomic status and the built environment on health.^7,8^ Parcel linkages require precise spatial alignment; unlike environmental exposures that often display smooth spatial gradients, parcel-level attributes change abruptly across property boundaries, magnifying the effect of linkage error and exposure misclassification. For example, a 20-meter displacement may move an address from a single-family home to a large apartment building, misclassifying land use type, assessed market value, and housing condition.

Integration of address data with parcel boundaries, building footprints, and other high-resolution environmental datasets allows researchers to refine exposure assignments to the level of individual residences,^9,10^ but this linkage can be technically challenging.^7^ Recent advances in natural language processing are improving address standardization and linkage accuracy. Open-source parsing tools such as Libpostal and usaddress help normalize and compare unstructured address strings.^11,12^ Further, the United States maintains several initiatives to improve the accuracy and consistency of address linkages and geocoding. The National Geospatial Advisory Committee identifies address data as critical public infrastructure,^13^ and the U.S. Department of Transportation coordinates the National Address Database (NAD) to integrate authoritative address points from state, local, and tribal sources.^14^ Using National Emergency Number Association and Federal Geographic Data Committee standards, the NAD provides a foundation for consistent geocoding across research, public health, and emergency response applications.

Despite this progress, address data present persistent challenges for research and clinical use. Formats and conventions vary across jurisdictions, and addresses can change over time through renumbering, annexation, or rural route conversion.^15^ Databases are often incomplete or inconsistently maintained, and the same address may produce different coordinates depending on the geocoding algorithm.^1,5^ Clinical data further complicate matters because patients may have multiple addresses, each with different implications for exposure or care. Matching addresses across datasets requires robust standardization and probabilistic linkage methods capable of handling errors, abbreviations, and typographical variation.

The impact of geocoding accuracy on bias and exposure misclassification in environmental epidemiology, particularly in air pollution studies, has been extensively documented.^16,17^ However, no comparable evaluations have been published examining how spatial inaccuracies affect linkage to address-level exposures, such as parcel- or property-based data, and in the literature, approaches to address-parcel linkage vary. Many studies rely on commercial geocoding software, such as ArcGIS, to match geocoded addresses to parcel boundaries or centroid points.^10,18–20^ Some researchers also supplement automated geocoding with manual inspection or rule-based correction to improve match quality.^6,21^ Without a clear understanding of how accurate different linkage methods are, the potential value of address data in health research is undermined, introducing uncertainty, bias, and inconsistency that limit the reliability of spatially derived findings or interventions. Identifying accurate and scalable address linkage strategies is critical as the use of precision population health approaches and the availability of address and parcel data grow.

Our objective was to evaluate address tag matching and geographic matching approaches for linking addresses in administrative and health records to parcel data with respect to the accuracy of parcel identifiers, parcel characteristics, and real-world address-level exposures.

## Methods

### Setting and data sources

To test linkage of addresses to parcels, we leveraged data from the NAD, a harmonized spatial database that contains address data, point location coordinates, and other supporting data for more than 30 states.^14^ NAD data originate from state authorities and are maintained through varied processes; municipal planning offices, emergency management agencies, and postal services each play a role in assigning, validating, and updating addresses. The methods used to assign geographic coordinates differ substantially; some rely on street range interpolation while others use parcel centroids or building footprint coordinates derived from cadastral data. This study focused on residential addresses in two urban and suburban counties in Ohio including Cincinnati (Hamilton County) and Columbus (Franklin County). In Ohio, NAD coordinates come from the Ohio Geographically Referenced Information Program’s Location Based Response System (LBRS). LBRS places points for all addressable locations along access from the road, typically on the structure’s driveway.^22^ Visual inspection indicated points are typically on or near the front of the building (Figure 3C).

Using addresses from the NAD, we used a gold standard linkage to parcel identifiers to determine the accuracy of different address- and geographic-based matching methods for linking parcel identifiers and accompanying parcel-level characteristics (Figure 1). The gold-standard relationship between residential addresses, property boundaries, and parcel attributes was based on authoritative geospatial databases from each county’s County or Area Geographic Information Systems (CAGIS; https://franklincountyauditor.com, https://hamiltoncountyauditor.org). For each county, an address-level dataset was used to link addresses to parcel identifiers, and a parcel-based dataset was used to link parcel characteristics via the parcel identifier. In addition to using the parcel boundary polygons, we calculated the parcel centroids using the Ohio South State Plane coordinate system (EPSG:3735). When using parcel-based datasets, we considered only residential parcels by filtering to those with relevant major land-use codes defined under the Ohio Administrative Code Rule 5703-25-10 (i.e., one-, two-, and three-family dwellings; condominiums, apartment buildings, and other commercial- or government-based housing).^23^ Address-based datasets were not similarly filtered because of a lack of an interoperable residential address type definition.

**Figure 1.**
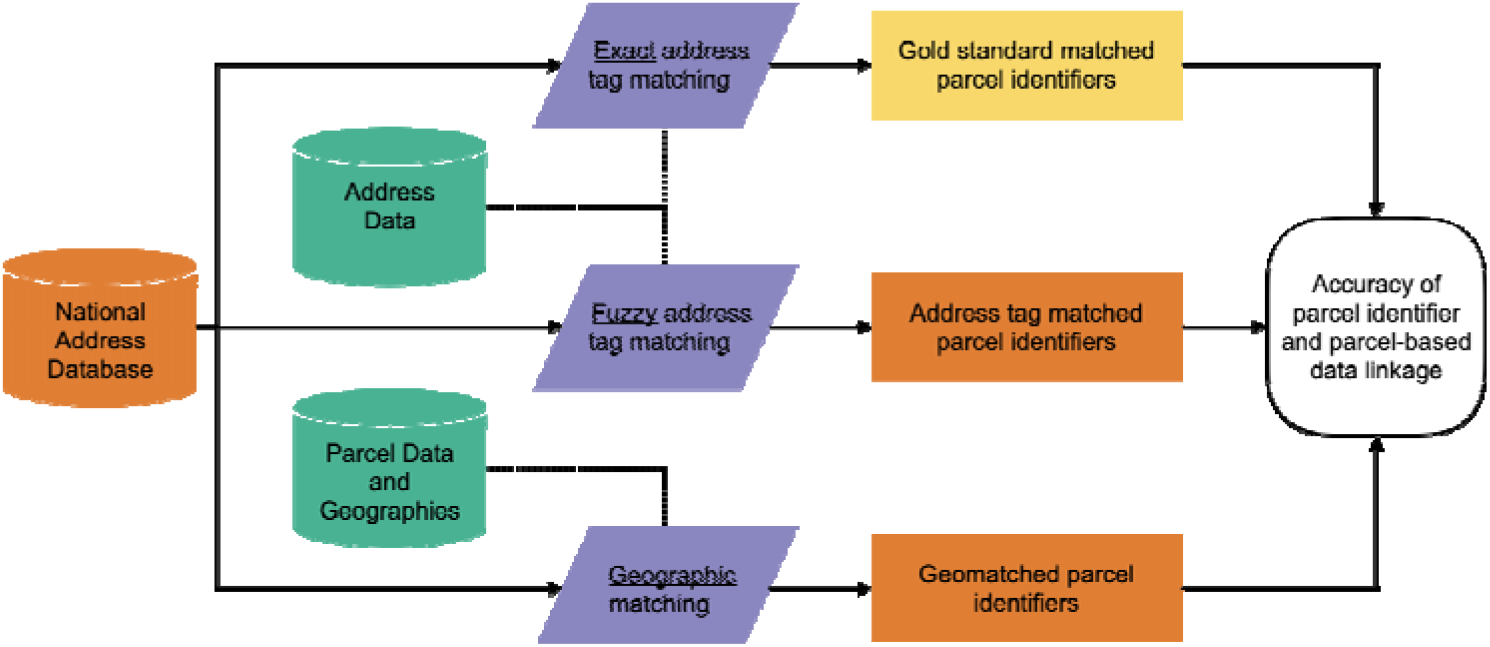
An overview of data and methods used to compare address to parcel linkage methods.

To estimate potential misclassification in a healthcare context, we used records from the Cincinnati Children’s Hospital Medical Center electronic health record. The data extract was done under an existing IRB protocol at Cincinnati Children’s that was classified as exempt from ongoing review. We extracted the most recent patient addresses from all demographics tables available in our instance of Epic (starting in 2010) on October 15^th^, 2025. Known institutional addresses (e.g., Ronald McDonald House, Hamilton County Job and Family Services) were excluded.

### Defining gold standard address-parcel linkage

To define the gold standard relationship between addresses in the NAD and addresses in the CAGIS address-parcel data, we parsed addresses into structured address components (i.e. “tags”) using version 0.7.0 of the addr package for R (https://github.com/geomarker-io/addr) and matched addresses exactly (string distance = 0 for street number, street name, street type, and ZIP code; Figure 1).^24^ We exactly matched 93.3% of Hamilton County NAD addresses and 77.0% of Franklin County NAD addresses to their parcel identifiers and excluded NAD addresses that did not match to any CAGIS addresses from further analysis. Manual inspection revealed that these were addresses no longer used in CAGIS sources, were highway addresses (e.g. “3900 I-275 WB Expressway”), or were addresses with street type modifiers that were unable to be correctly tagged (e.g. “999 LOST Crossing 3-H”, “3481 Amerway court 92”). Some NAD addresses matched to duplicated addresses in CAGIS data, which occurred if addresses matched on all tags except for placename and city state (e.g., “6861 Rapid Run Road Delhi Township OH 45233” and “6861 Rapid Run Road Cincinnati OH 45233”), or CAGIS addresses associated with multiple parcels; in these cases we considered the alphabetically-sorted first address to be the gold standard match.

### Matching approaches

Addresses can be matched using either (1) address tag matching: string matching of specific address components, like street name, number, or ZIP code; or (2) geomatching: geographic matching of geocoded addresses through intersection with parcel boundaries or distance to parcel centroid (Figure 2; Figure 3). Both can rely on natural language processing to identify structured components from messy, real-world residential addresses, but address tag matching provides the specificity of address-level linkage that geocoders are not designed to provide. Geomatching relies on minimizing the calculated physical distance between two geocoded coordinates or testing for geographic intersection between a point and polygon, both of which may vary based on the geocoding method itself, the algorithm used to calculate spatial distance and intersection, and the spatial projection used to complete the calculations. Address tag matching allows for tag-specific thresholds (e.g., one string edit difference for street name vs. exact match for street number), which are more appropriate for linkage of address-level exposures and characteristics.

**Figure 2.**
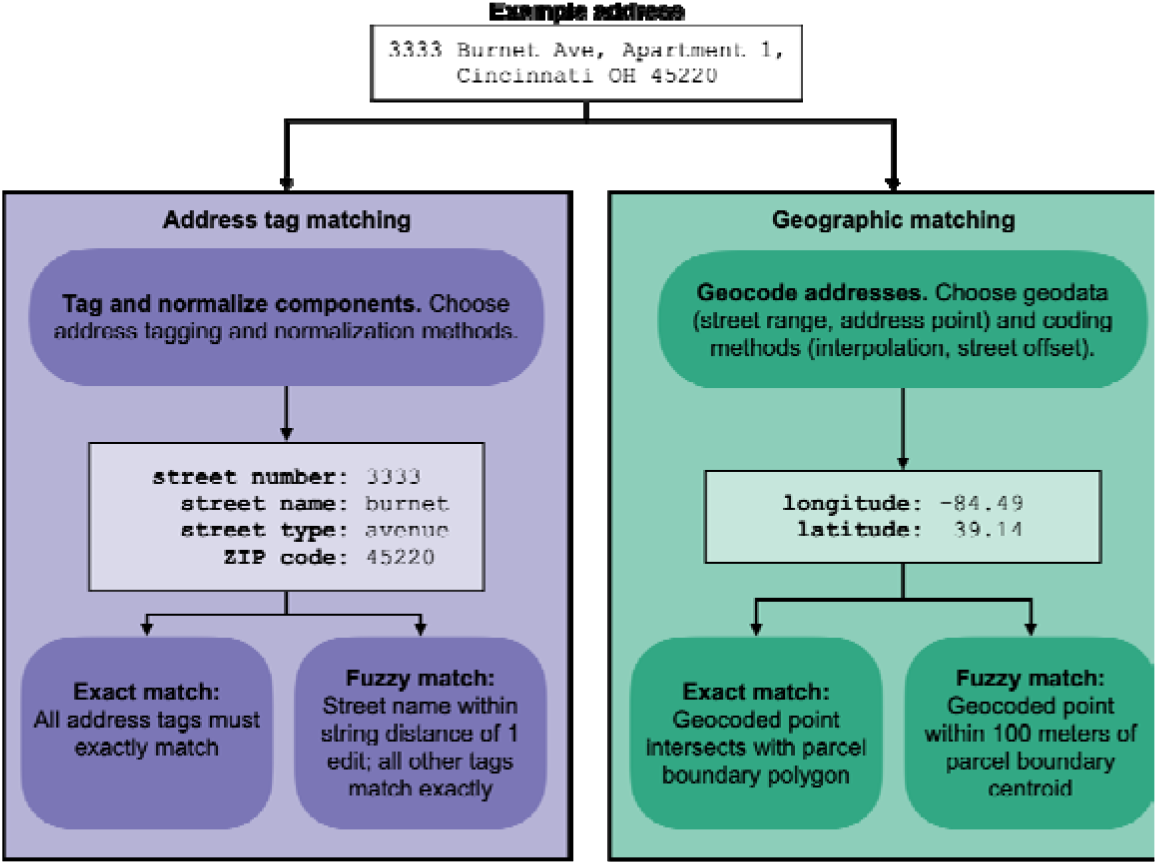
A comparison of address tag and geographic matching workflows.

We attempted to match Hamilton and Franklin County, Ohio, addresses in the NAD to their gold-standard addresses and parcel identifiers from their respective CAGIS using traditional geocoding-based approaches (geomatching) and a deterministic, text-based address linkage method (address tag matching; Figure 2).^7^ For geomatching, we used the NAD coordinates (derived the Ohio Geographically Referenced Information Program’s Location Based Response System) for each address as its address point geocoded coordinates (Figure 3C). We also geocoded NAD addresses with a street range geocoder (DeGAUSS^25^) to determine the impact of geocoding precision on matching performance (Figure 3D). We geomatched by choosing the nearest parcel centroid within 100 meters or by geographic intersection with the parcel boundaries (Figure 3A-B) using the Ohio South State Plane coordinate system (EPSG:3735). For address tag matching, we used addr R package (v0.7.0) to parse addresses into tags and directly link NAD addresses with CAGIS address records with fuzzy string matching (string distance = 1 for street name; string distance = 0 for street number, street type, and ZIP code).^24^

**Figure 3.**
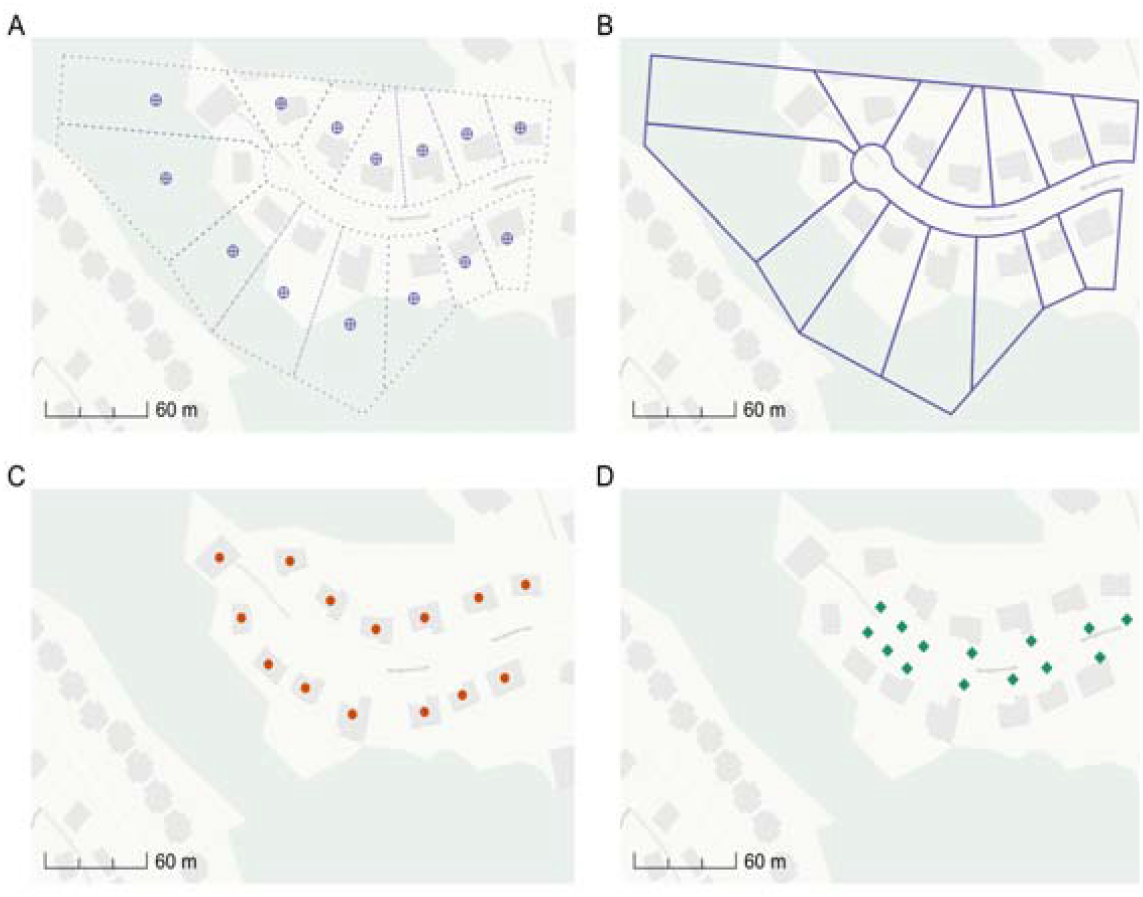
Maps of A) parcel centroids, B) parcel polygons, C) address point coordinates, and D) street range geocoded coordinates for a representative street in Hamilton County, Ohio.

### Matching performance evaluation

For each county, we calculated the proportion of NAD addresses that were successfully matched to a parcel, defined as having a single, non-missing parcel ID. Address tag matching and geomatching with parcel polygons can result in one address matching multiple parcels. Multi-matches in address tag matching were resolved to the first alphabetical match since they were all identically matched, but we retained multi-matches for polygons since they all intersected and did not consider these addresses for parcel attribute agreement. Agreement across matching methods was evaluated relative to the gold standard. We calculated parcel ID agreement across matching methods as well as agreement in parcel-level attributes, land use type (exact match) and market total value (within 20%), as one metric. Parcel-level attributes were only linked for residential parcels; others were considered missing. In both cases, two missing values were considered agreeable.

To assess the impact of matching method on linkage of a real-world parcel-level exposure (poor housing quality), we used open data from the City of Columbus on enforced housing code violations between January 1, 2019, and October 15, 2025, which include an address and parcel ID for each record.^26^ We calculated sensitivity for each method based on parcel ID agreement.

To examine how hyperlocal address density impacts matching performance, we first counted the number of NAD address points within 100 meters of each address’s NAD coordinates. We then assigned addresses to 2020 census tract geographies and calculated each tract’s average hyperlocal address density. Finally, we stratified tracts into quartiles based on average density and calculated land use and value agreement within each; we used Kruskall-Wallis tests to evaluate differences between quartiles for each method and county. To assess for equity implications, we also stratified tracts into quartiles based on a community material deprivation index^27^ and evaluated their agreement with the address density quartile classifications using a chi-squared test for each county.

To estimate the impact of different methods when linking real-world healthcare addresses, we evaluated matching performance when weighting each address by the number of patients in the Cincinnati Children’s electronic health record.

Finally, we assessed the runtime of the addr package (version 0.9.9) on a laptop equipped with Apple M2 Pro (10-core CPU, 16 GB RAM), applying fuzzy tag matching to link randomly selected addresses of Hamilton County registered voters^28^ to the National Address Database for Hamilton County, Ohio.

## Results

We defined the gold standard match between distinct NAD addresses in Hamilton (n = 332,988) and Franklin Counties (n = 559,497), Ohio, and distinct CAGIS address-parcel pairs with non-missing street number, street name, zip code, and parcel identifier in Hamilton (n = 347,174) and Franklin Counties (n = 686,806), resulting in 853,255 addresses from both counties linked to a gold standard parcel identifier that we used to test the different address-parcel matching strategies. To obtain coordinates for geomatching, all addresses had address point coordinates from the NAD, and 97.4% of addresses were geocoded using a street range geocoder.

Overall, availability of matched addresses and their agreement with the gold standard varied across matching methods and between the two counties (Table 1). Geomatching to parcel centroids returned at least one match candidate more often than geomatching to parcel polygons (e.g., 96.5% versus 35.1% in Hamilton County when using range-based geomatching; Table 1). Across address point and street range geomatching methods in both counties, nearly all addresses were geomatched to parcel centroids (87.5% - 97.2%), but geomatching to parcel polygons matched much less often (26.1% - 82.8%; Table 1).

**Table 1.**
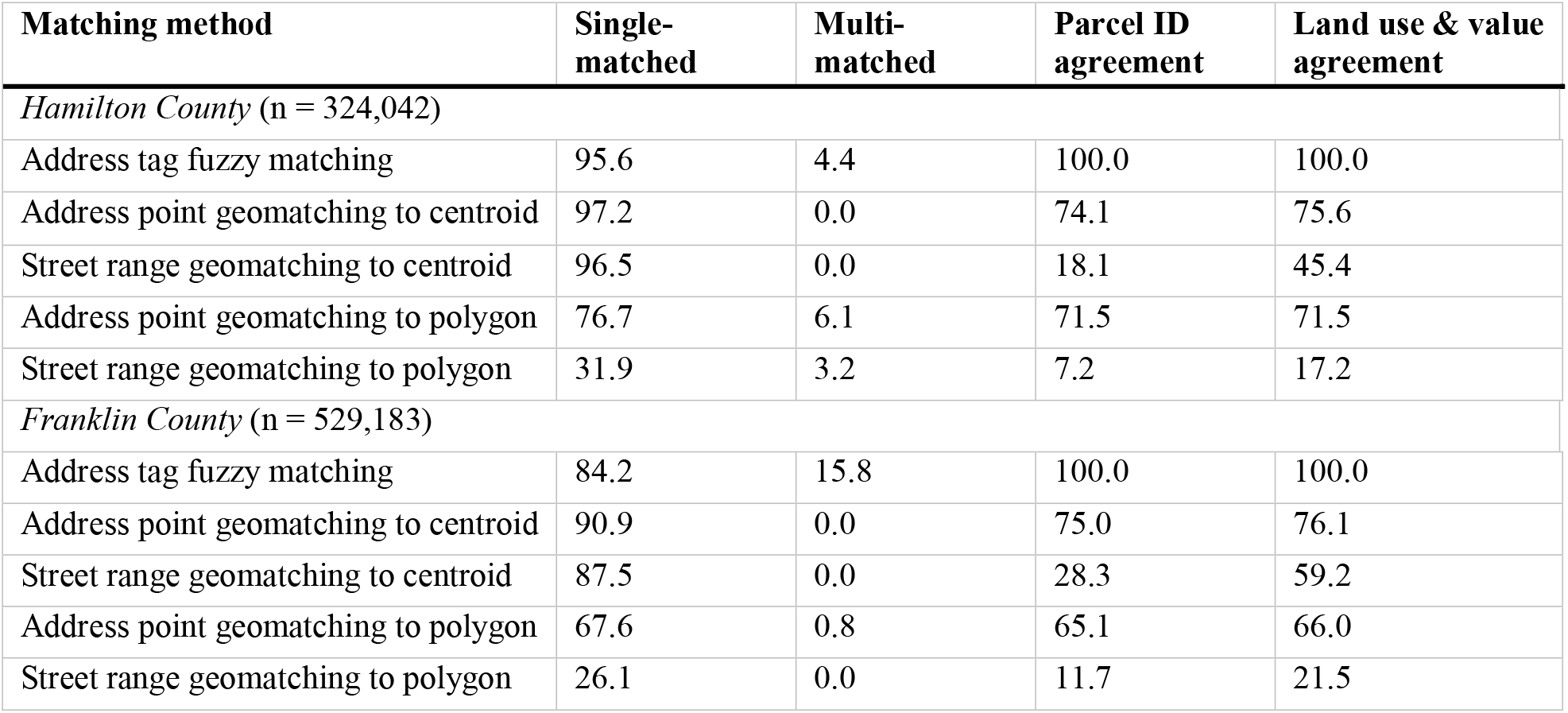
Matching evaluation metrics (%) by method and county.

In the case of geomatching using parcel polygons, multiple matches occurred more often in Hamilton County (3.2% - 6.1%) compared to Franklin County (0.0% - 0.8%; Table 1). The median number of parcels per multi-matched address ranged from 4 (address point geomatching to polygons in Franklin County) to 17 (address point geomatching to polygons in Hamilton County). The majority of parcels associated with multi-matched addresses were condominiums, with the exception of address tag matching in Franklin County, where the most common land use type was 40+ unit apartments.

Among all geomatching methods, address point geomatching to parcel centroids had the highest agreement with the gold standard (74.1% - 76.1% for both ID and land use/value; Table 1). In contrast, street range geomatching consistently had the lowest agreement (7.2% - 28.3% for parcel ID and 17.2% - 59.2% for land use/value; Table 1). Address tag fuzzy matching resulted in multiple matched addresses in Hamilton County (4.4%) and Franklin County (15.8%; Table 1). In these cases where one address is associated with more than one parcel (e.g., condominiums), the parcel identifiers and land use/values agreed for all matches; address tag fuzzy matched parcel identifiers as well as land use and value agreed 100% with the gold standard, performing better than any of the geomatching methods.

We linked all 38,457 enforced housing code violations associated with 22,248 parcels to 7.8% of Franklin County NAD addresses (n = 53,904) based on exact address tag matching. The sensitivity, based on parcel ID agreement, was highest using address tag fuzzy matching (100%) and address point geomatching (97.5% and 97.1% using centroids and polygons, respectively), indicating strong performance for linkage of real-world parcel-level exposures. Street range geomatching to polygons exhibited the lowest sensitivity at 57.0%.

We found differences in tract-level land use and value agreement across quartiles of hyperlocal address density for all matching methods in both counties (Kruskall-Wallis test p < 0.001; Table 2). In general, street range geomatching approaches had lower agreement than address point geomatching, but increasing hyperlocal address density was associated with decreasing agreement in all methods (Table 2). We observed significant agreement between address density- and community deprivation-based quartiles in both counties (Chi-squared test p < 0.001).

**Table 2.** Land use and value agreement (%) for geomatching methods, by tract-level hyperlocal address density quartile and county. Address tag matching had 100% agreement across quartiles.

| **Address density quartile** | **Mean number of addresses within 100m** | **Address point geomatching to centroid** | **Street range geomatching to centroid** | **Address point geomatching to polygon** | **Street range geomatching to polygon** |
| --- | --- | --- | --- | --- | --- |
| *Hamilton County (n = 324,042)* |  |  |  |  |  |
| 1 | 5.6 - 23.2 | 80.8 | 54.1 | 78.0 | 22.9 |
| 2 | 23.2 - 30.7 | 81.9 | 52.7 | 78.8 | 20.1 |
| 3 | 31.0 - 42.2 | 70.7 | 34.5 | 67.0 | 11.6 |
| 4 | 42.3 - 169.0 | 58.0 | 25.5 | 50.6 | 6.8 |
| *Franklin County (n = 529,183)* |  |  |  |  |  |
| 1 | 7.5 - 34.5 | 82.4 | 65.0 | 79.8 | 22.1 |
| 2 | 34.7 - 51.8 | 84.9 | 68.2 | 78.9 | 24.5 |
| 3 | 51.8 - 72.6 | 76.0 | 55.9 | 65.8 | 21.7 |
| 4 | 72.6 - 290.0 | 68.2 | 50.4 | 46.0 | 16.0 |

We extracted 792,855 unique addresses associated with 1,359,125 patients from the Cincinnati Children’s electronic health record and linked 144,987 of them (representing 429,317 patients) to Hamilton County NAD addresses (41.8% of all addresses). When calculating evaluation metrics with weighting by the number of patients, the percentage of multi-matched addresses was higher for address tag fuzzy matching and geomatching to polygons methods (e.g., address point geomatching to polygons resulted in multiple matches for 25.8% of addresses, compared to 6.1% of all addresses in Hamilton County), suggesting that more people live at addresses associated with multiple parcels (e.g., large, multi-unit buildings; Table 3). Parcel ID and land use/value agreement for geomatching to polygons methods was lower (e.g., 65.2% of addresses matched by address point geomatching to polygons had parcel ID agreement vs. 71.5% of all Hamilton County addresses), indicating that linkage disagreements are magnified when considering real-world frequency of address usage in the electronic health record (Table 3).

**Table 3.** Matching evaluation metrics (%) for Hamilton County addresses found in the Cincinnati Children’s electronic health record (n = 144,987), weighted by the number of patients per address.

| Matching method | Single-matched | Multi-matched | Parcel ID agreement | Land use & value agreement |
| --- | --- | --- | --- | --- |
| Address tag fuzzy matching | 74.5 | 25.5 | 100.0 | 100.0 |
| Address point geomatching to centroid | 99.6 | 0.0 | 83.5 | 80.9 |
| Street range geomatching to centroid | 99.3 | 0.0 | 24.4 | 48.5 |
| Address point geomatching to polygon | 71.3 | 25.8 | 65.2 | 65.1 |
| Street range geomatching to polygon | 27.6 | 4.7 | 6.9 | 15.5 |

In run time testing of our address tag fuzzy matching pipeline, we processed 10,000 addresses spanning 54 ZIP codes in 2 minutes, 14 seconds, and 100,000 addresses spanning 55 ZIP codes in 19 minutes, 50 seconds.

## Discussion

Different approaches to linking addresses to parcels produce widely varying results, indicating that results of studies using address-parcel linkage may be sensitive to their method choices. Understanding the strengths and limitations of each approach and the acceptable level of geospatial uncertainty for the study design and exposure measurement is essential for selecting an optimal linkage method.^29^

Overall, geomatching methods were sensitive but showed limited specificity, producing high rates of non-missingness but poor agreement with the gold standard. Street range geomatching to parcel polygons was an exception, with much lower completeness and very low agreement. Address point geomatching offered the best balance of completeness and accuracy among geographic-based methods, although even these approaches produced exposure misclassification rates of 20–35%, levels that are generally not acceptable for parcel-level exposure assessment. Importantly, parcel polygons are not universally available, and spatial intersection methods can be computationally intensive at scale and require thoughtful selection of geospatial projections and computations.

In contrast, address tag fuzzy matching, which directly matches parsed address components to CAGIS records using open-source tools, achieved 100% accuracy and 0% missingness. While some agreement with the gold standard is likely due to their common address tagging and normalization methods, these choices are indeed the major advantages of a fuzzy matching method where fine control over multi-variate matching distances provides a more customizable and useful tool to link address-level data. Further, address component specificity avoids the spatial uncertainty introduced by geocoding, so this method may be preferable for high-resolution analyses where small positional errors can substantially alter exposure estimates.

Match rates remain a challenge in real-world studies using parcel-level exposures. For example, Wilson et al. matched 60.5% of patient addresses from asthma encounters to parcels using a multi-stage geomatching approach,^6^ and Laurent et al. matched 54.0% of birth certificate addresses by parcel polygon intersection.^20^ In contrast, Shi et al. were able to match 87.1% of physician practice addresses using an address tag matching approach.^15^ These prior findings are consistent with our results, emphasizing the value of robust tag-based linkage strategies for achieving accuracy and completeness in parcel-level exposure assessment.

The incidence of multi-matches is a key limitation to using address tag matching and polygon intersection approaches. Condominiums, apartments, and other multi-unit residential complexes present a challenge in which many parcels and many addresses may be legitimately associated with a single building or property. For example, “2444 MADISON Road 209 CINCINNATI OH 45208”, associated with a unit in a large condominium building, is parsed to “2444 madison road 45208” and matched to 259 unique parcel IDs. This is consistent with findings by Zandbergen that address point geocoding was unreliable for multi-unit residential addresses.^5^ There are no established best practices for resolving multiple matches. When using address tag matching, we resolved multi-matches by reproducibly selecting one, but all matched identifiers could be compared in cases where one-to-many or many-to-many linkages are expected. Geomatching by intersecting geocoded points to parcel polygons can also produce multi-matches when a point intersected with more than one parcel boundary, but they were not resolved to a single match. Additional research could assess the validity of summarizing parcel attributes across matched parcels (e.g., mode of land use, mean or sum of market value) or developing more precise methods to incorporate “Line 2” of addresses (e.g., unit numbers) or the vertical dimension of polygons for multi-story buildings. Hybrid or staged matching strategies, such as Wilson et al.’s multi-phase workflow involving range-based geocoding, address point geocoding, and manual inspection, may offer further improvements.^6^

The performance of address tag matching relies heavily on the availability, quality, and completeness of address-parcel datasets on which to base a gold standard, and this varies considerably by municipality.^5,30^ Substantial variability in our results between Hamilton and Franklin Counties shows that local data governance, including address maintenance practices and parcel dataset quality, can impact matching accuracy. Increasing data harmonization and standardization efforts could improve the robustness and scalability of address tag-matching methods by reducing cross-jurisdictional variability. Existing examples include commercial sources like Regrid (https://regrid.com/), which hosts nationwide harmonized parcel data, and states that maintain large, standardized datasets, such as Massachusetts’ MassGIS and Indiana’s GIS Data Harvest.^31,32^ The NAD and Next Generation 9-1-1 represent similar efforts at the national level.^14,33^

Restricting the target parcel records to only those that are residential increases the match quality for input residential addresses, but addresses in open datasets may not use a standard categorization or ontology for type or land use. For example, Hamilton and Franklin Counties are both in Ohio and use land use categories defined in the Ohio Administrative Code in the parcel geographies but different mappings for address type. This again emphasizes the importance of regional or national standardization efforts.

Hyperlocal address density was strongly associated with matching performance, and the association between address density and community deprivation raises equity concerns. Errors are more likely in multi-family buildings and dense urban neighborhoods, which disproportionately house socioeconomically disadvantaged communities. This pattern is consistent with Kinnee et al.’s findings of greater air pollution exposure misclassification in low socioeconomic status areas, although they did not see differences in geocoding distance error by ZIP code-level address density.^16^ Similarly, Brooks et al. found agreement between participant addresses in a national longitudinal study and a commercial address database varied by sociodemographic groups.^1^

We performed this analysis in two predominantly urban and suburban counties in one state. As such, findings may not be generalizable to more rural areas, where infrastructural constraints, addressing conventions, and data completeness challenges may affect performance. However, address tag matching and address point geomatching are still likely to outperform traditional street range geocoding approaches. Similarly, regions with different data governance structures and maintenance practices may experience different levels of linkage accuracy. Broader evaluations across jurisdictions would help clarify the external validity of our findings, but this relies on data harmonization feasibility. The NAD is available in most U.S. states, making this approach broadly implementable where address-parcel datasets are accessible. Successful application requires careful consideration of the specific local data sources, as we have demonstrated in our study.

Integrating address tag matching pipelines into large healthcare systems or public health surveillance infrastructure requires attention to computational efficiency, data management, and routine data quality checks. The addr R package can be integrated into existing workflows as a modular preprocessing layer that standardizes, parses, and probabilistically links messy real-world address data prior to geocoding, spatial linkage, and analytic modeling, thereby improving data quality, reproducibility, and scalability within routine surveillance and research pipelines. Unlike conventional high-precision geocoding, which depends on external services and point-level coordinate assignment, addr operates directly on structured address components, enabling deterministic linkage, transparent error handling, reduced privacy risk, and greater control within secure computing environments. Run time testing indicated modest but nontrivial computational demands that scale approximately linearly with input size and remain feasible for large-scale clinical databases given contemporary workstation-class hardware.

## Conclusion

Policymakers and health agencies can support these efforts by promoting standardized address maintenance practices, incentivizing open data availability, and developing regulatory guidance for geospatial data linkage used in health research. Parcel data have already proved valuable in studies related to lead poisoning risk, the impact of housing code violations on asthma, and address-level pediatric hospitalization risk.^8,9,33^ Improving address-parcel linkage methods will enhance the accuracy of such applications, enabling more precise and reliable environmental exposure assessments. Future research should examine how differences in address-parcel linkage propagate through epidemiologic analyses, including bias in exposure metrics, effect size estimates, and health outcome associations.

## Data Availability

All data used in this study, with the exception of patient addresses from healthcare records, are publicly available online.

https://github.com/geomarker-io/addr

https://github.com/geomarker-io/parcel

https://apps.franklincountyauditor.com/

https://opendata.columbus.gov/datasets/columbus::code-enforcement-cases/about

https://cagis.org/Opendata/

## Acknowledgements

Research reported in this publication was supported by the National Institute of Environmental Health Sciences of the National Institutes of Health under Award Number R03ES037996. The content is solely the responsibility of the authors and does not necessarily represent the official views of the National Institutes of Health. We gratefully acknowledge the many county agencies and regional GIS consortiums whose commitment to open geospatial data make this work possible.

